# Application of 7-1-7 framework in prototypical anthrax outbreak: Identifying missed opportunities for early detection in Southwestern Uganda, September 2024

**DOI:** 10.1101/2025.09.23.25336512

**Authors:** Hannington Katumba, Richard Migisha, Charity Mutesi, Bridget Ainembabazi, Loryndah Olive Namakula, Hellen Nelly Naiga, Aloysius Tumwesigye, Birungi Mutahunga Rwamatware, Hilda Tendo Nansikombi, Lilian Bulage, Benon Kwesiga, Milton Bahati, Martin Esau, Paul Johnson Lumu, Gladys Kiggundu Nakanjako, Stella Acaye Atim, Sarah Acayo, Linda A. Atiku, Julius J. Lutwama, Pontiano Kaleebu, Lydia Nakiire, Dathan M. Byonanebye, Francis Kakooza, Alex Riolexus Ario

## Abstract

**Introduction:** Uganda responded to 37 anthrax outbreaks during 2014–2024. In 2021, the country adopted the 7-1-7 framework, which stipulates detection of outbreaks within seven days, notification completed in one day, and completion of early response actions initiated within seven days. However, the application of the 7-1-7 timeliness evaluation for prototypical zoonotic infections, such as anthrax, is unclear. We assessed the timeliness of response to the 2021 Anthrax outbreak in a rural border district in Uganda.

**Methods:** We reviewed human and animal health surveillance records to document key dates of emergence, detection, notification, and completion of early response actions. We used the 7-1-7 metrics to evaluate the timeliness of these milestones. Through discussions with district leadership, health workers, and residents, we verified key dates and facts, identified bottlenecks and enablers, and confirmed the accuracy of information. Qualitative data were organized into themes to capture bottlenecks and enablers.

**Results:** The disease in animals was detected after 87 days, and notification was made in one day. In humans, detection took 83 days, and notification took one day. Early response actions were jointly initiated five days after notification and completed within 10 days. The outbreak involved 111 animal deaths and 90 cases of human anthrax, including 6 (6.7%) deaths. The enablers included the presence of a real-time One Health communication platform, partner support, presence of Village Health Teams (VHTs), and training of animal health workers on detection. Bottlenecks included weak zoonotic disease surveillance, characterized by understaffing, a low suspicion index, misdiagnosis, and weak coordination with private health facilities.

**Conclusion:** Overall, there were delays in detection and completion of early response in an anthrax outbreak in humans and animals, highlighting the need for cross-sectoral coordination to ensure coordinated surveillance and response in a One Health approach. Strengthening animal health surveillance, building private sector engagement, and training frontline workers to recognise zoonoses early could improve future outbreak responsiveness and mitigate potential cross-border spills.

## INTRODUCTION

Timely detection and response to public health threats is an integral component of the International Health Regulations (IHR) [1, 2]. With increased global connectivity, the risk of emerging and re-emerging infectious diseases, as well as other public health threats, spreading across national boundaries increases. This is a reality that emphasizes the critical need for effective and robust surveillance systems that can prepare for, detect, and respond promptly to such threats. According to the Global Health Security Index (GHSI) of 2021, with an average preparedness score of 38.9%, there was no country fully prepared for a pandemic [3].

There is growing evidence that most high-impact outbreaks in recent decades, such as COVID-19, Ebola, SARS, MERS, and mpox, have all been zoonotic, highlighting the need for coordination of surveillance among the animal, human, and environmental health sectors worldwide. Factors contributing to zoonotic spillovers, such as urban expansion, climate change, and increased population mobility, are becoming more pronounced, particularly in sub-Saharan Africa (SSA) and other low- and middle-income regions. This leads to increased frequency and complexity of infectious disease outbreaks, further highlighting the need for integrated and multi-sectoral surveillance and response approaches guided by the One Health framework.

Through the IHR, the World Health Organization (WHO) makes it imperative for all national governments to strengthen their capacities to detect, assess, and respond to public health threats of international concern. As a result, countries need to implement rapid disease prevention and control measures [4]. Of particular concern are countries in sub-Saharan Africa, where, amidst rising human populations, countries have experienced numerous public health emergencies in recent years, including disease outbreaks and natural disasters [5, 6]. An increased human population often comes with resultant improved standards of living in urban communities and increased demand for animal products [7, 8]. This dynamic provides impetus for increased engagement in livestock rearing in rural communities, where this activity is more suitable. As a result, the livestock population in Uganda has shown an annual increasing trend, with a subsequent increase in the interaction between humans, animals, and the environment [9]. Given projected population growth in many SSA countries and the associated pressure on public health systems, strengthening surveillance systems is essential to ensure timely outbreak response and minimise impacts on individuals, communities, and the economy, and ultimately fulfil IHR core capacities. In this context, the speed of case detection remains a critical measure of surveillance system effectiveness, particularly for priority diseases [10, 11].

The WHO recommends the 7-1-7 framework for assessing the timeliness of response to public health emergencies. According to this framework, public health emergencies should be detected within 7 days from the date of emergence, a notification made to the authority responsible for action within one day, and seven early response actions completed within seven days after notification. The WHO African region and international donor agencies have adopted the 7-1-7 framework. Additionally, several national governments, including Uganda under the Ministry of Health, have adopted and are currently applying the 7-1-7 metrics on different outbreaks such as cholera, measles, and anthrax [12, 13].

Anthrax, particularly, is a fatal zoonotic disease caused by *Bacillus anthracis*, a resilient spore-forming bacterium. The organism is naturally resident in the soil and can survive in the environment for decades, making it quite challenging to eliminate once introduced. Anthrax spores can be activated under favourable conditions, such as heavy rains with floods or during droughts, where livestock graze on nearly bare ground. In such instances, animals become exposed by feeding on spores in the soil or contaminated water, and humans are exposed through contact with sick or dead animals or by consuming animal products from these animals. The incubation period differs by form of anthrax, ranging from 1 to 14 days in animals and up to 6 weeks in humans [1, 14]. The disease is a substantial threat to both public health and animal production and trade. In livestock, outbreaks can cause high mortality rates, leading to significant economic losses, disruption of meat and livestock trade, and long-term impacts on pastoralist and farming livelihoods. In humans, cutaneous anthrax can be fatal without treatment, while inhalational and gastrointestinal forms carry even higher case fatality rates. Furthermore, due to its stability, ease of dissemination, and high lethality, *Bacillus anthracis* is classified as a Category A bioterrorism agent by the U.S. Centers for Disease Control and Prevention. These factors underscore why anthrax remains a priority zoonotic disease in many SSA countries [11, 15-21].

Given the persistence and high impact of zoonotic outbreaks like anthrax, structured evaluation frameworks are essential for strengthening outbreak preparedness and response. The 7-1-7 target provides a clear benchmark: detect outbreaks within 7 days, notify within 1 day, and complete early response actions within 7 days. This approach allows countries to assess real-time performance during an outbreak and uncover operational gaps that may not be evident through routine reporting and public health service delivery. When done early in an outbreak as an Early Action Review (EAR), the 7-1-7 assessment allows identification of short-term bottlenecks, which can then be addressed. Additionally, the 7-1-7 model supports intra- and after-action reviews (IARs and AARs) by providing specific timelines that help identify systemic delays, inefficiencies, or coordination challenges across sectors. It can serve as a tool for evaluating outbreak response by focusing on the timeliness of interventions, a critical factor in zoonotic events like anthrax, where early action can prevent widespread transmission and economic loss. Furthermore, the framework enables quantifiable measurement of surveillance and response system performance, helping governments and partners to track progress, compare outcomes across geographical regions and sectors, and guide targeted improvements. The data generated through 7-1-7 assessments can also be used to inform advocacy for resource mobilisation and long-term policy reform, especially where repeated bottlenecks reveal structural weaknesses. By highlighting where delays consistently occur, whether in frontline detection, reporting, or cross-sectoral coordination, countries can make evidence-based decisions for system improvement. Within a One Health context, 7-1-7 offers a practical framework for monitoring and strengthening multisectoral preparedness, as well as shaping political commitment and budgetary prioritization. The application of this framework has been evaluated and documented in various countries, including Uganda [12, 22]. We applied this 7-1-7 framework to the first known anthrax outbreak in Kanungu District in Southwestern Uganda in September 2024 to assess the timeliness of detection, notification, and response, and identified the bottlenecks and enablers, to inform this and future similar responses.

## METHODS

### Outbreak setting

Kanungu District is located in Kigezi region, Southwestern Uganda. It is bordered by Rukungiri District in the east, Rubanda and Kisoro Districts in the south and by the Democratic Republic of Congo (DRC) in the west. It is also home to some sections of the Bwindi Impenetrable National Park (BINP) in the south and the Ishasha sector of the Queen Elizabeth National Park (QENP) in the North and North East (Figure 1).

**Figure 1.**
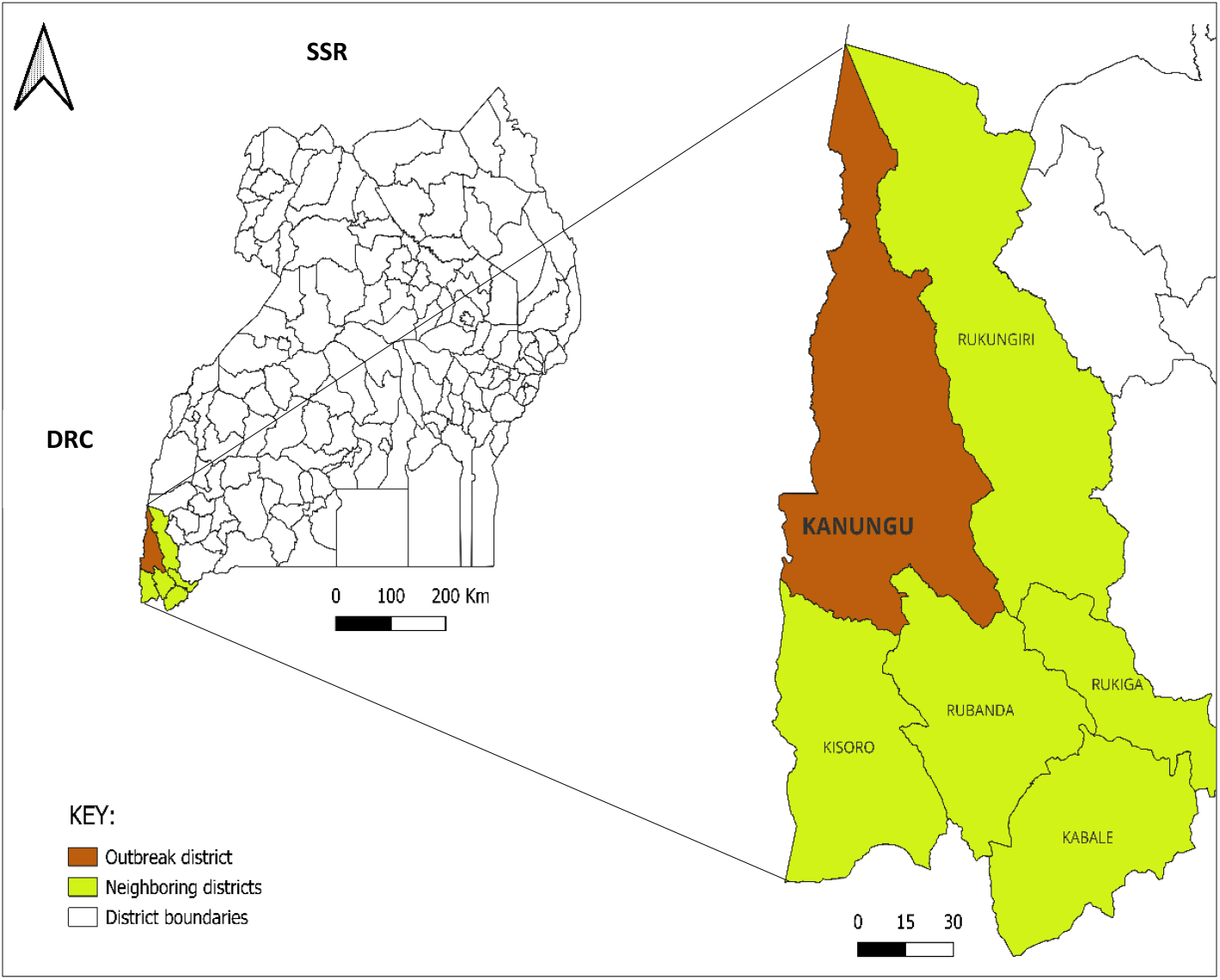
Location of Kanungu District relative to other districts and the Democratic Republic of Congo (DRC), October 2024; DRC=Democratic Republic of Congo; SSD= South Sudan Republic.

It is an agro-pastoral area with several livestock farms and communal grazing areas, and is a gateway for the movement of livestock and livestock products to the DRC. The outbreak in this district came following reports of sudden deaths of cattle, which were considered to be due to Black Quarter, a clostridial disease caused by *Clostridium chauvoei, and* also causes sudden death in seemingly healthy livestock. Human, animal and environmental samples were collected, and an outbreak of anthrax in humans was confirmed. The Uganda Ministry of Health (MoH) deployed the National Rapid Response team (NRRT) to support the district in the response. In this outbreak, 111 livestock (cattle and goats) died, and 90 cases of human anthrax were identified, including six deaths. The district had a District One Health Team (DOHT), comprising technical staff from the health department, veterinary sector, administration and political leadership.

### Data collection

To establish key dates and events, we reviewed records at the district veterinary office and the district health office. More records were reviewed at health facilities that reported cases, as well as subcounty veterinary offices. We documented the dates of emergence in both animals and humans, as well as the dates the outbreak was detected and when the district health authorities were notified. Additionally, we documented the dates of initiation and completion of the seven early response actions (Table 1). We then conducted interviews with health workers, field veterinarians, community members, and district-level technical, political and administrative leadership to identify the bottlenecks and enablers in the outbreak response and additionally corroborate the information obtained from the field.

**Table 1:** The 7-1-7 timeliness milestones and their definitions.

| Milestone | Definition |
| --- | --- |
| **Date of emergence** | For endemic diseases, it is the date on which a predetermined increase in case incidence over baseline rates occurred<br>For non-endemic diseases, the date on which the index case or first epidemiologically linked case first experienced symptoms<br>In this outbreak, the emergence in animal health was considered to be the date of the earliest linked animal death<br>For other public health events, date the threat first met criteria as a reportable event based on the country reporting standards |
| **Date of detection:** | Date the event is first recorded by any source or in any system. In this setting, detection in humans was when suspected case-patients were recorded at Mburameizi HC III. In animal health, detection was when a sudden animal death with suspected anthrax was reported to the subcounty animal health worker. |
| **Date of notification:** | Date the event is first reported to a public health authority responsible for action. Notification in this outbreak was when suspected human and animal anthrax were first reported to the District Health Officer (DHO) and the District Veterinary Officer (DVO). |
| **Date of completion of early response actions** | Date on which all applicable early response actions were completed. The following are the early response actions:<br>1. Initiate an investigation or deploy an investigation or response team<br>2. Conduct epidemiological analysis of burden, severity, and risk factors, and perform initial risk assessment |
|  | 3. Obtain laboratory confirmation of the outbreak aetiology<br>4. Initiate appropriate case management and infection prevention and control measures in health facilities<br>5. Initiate appropriate public health countermeasures in affected communities<br>6. Initiate appropriate risk communication and community engagement activities<br>7. Establish a coordination mechanism |

### Quantitative data analysis

Based on the above definitions, the time to detection was determined as the time in days obtained by subtracting the date of emergence from the date of detection. The time to notification was calculated as the number of days between the detection date and the date when the DVO and DHO were notified. The time to response completion was the time from the date of notification to the date of completion of the seven early response actions.

### Qualitative analysis

We used thematic analysis for the qualitative study; Three (3) members of the investigation team, proficient in the local language (Runyankole-Rukiga), listened to audio recordings and analysed the information in each sentence. The data were manually coded and grouped thematically into bottlenecks and enablers as highlighted by the respondents. Any disputes were resolved by consensus. We presented the findings and quoted verbatim some representative statements obtained from respondents, highlighting the bottlenecks and enablers for detection, notification and response in animal and human health.

### Ethical approval and consent to participate

We conducted this study in response to a public health emergency, and as such, it was determined to be non-research. The MoH authorised this study, and the office of the Center for Global Health, US Center for Disease Control and Prevention determined that this activity was not human subject research, with its primary intent being for public health practice or disease control. This activity was reviewed by CDC and was conducted consistent with applicable federal law and CDC policy. §§See e.g., 45 C.F.R. part 46, 21 C.F.R. part 56; 42 U.S.C. §241(d); 5 U.S.C. §552a; 44 U.S.C. §3501 et seq.

We obtained additional permission to investigate from the Kanungu District local government health authorities. We obtained verbal consent from all the respondents aged ≥ 18 years, and there were no respondents under 18 years of age. Participants were assured that their participation was voluntary and that there would be no negative consequences for declining to participate or withdrawing from the investigation. Data collected did not contain any individual identifiers, and information was stored in password-protected computers, only accessible to the investigation team.

## RESULTS

### Time characteristics of the outbreak

#### Animal health

Sudden deaths of eight bulls were reported on animal Farm A from June 15-June 22, 2024. All the dead animals on this farm were slaughtered, with the meat distributed to community members, butcher owners, and traders dealing in meat from dead animals.

As animal deaths continued across the district, sudden cattle deaths occurred in Farm B as well and was attributed to Black Quarter disease. Farm B and other farms across the district then embarked on vaccination against Black Quarter disease.

On September 9, 2024, following detection of anthrax in humans at a health facility, a notification of suspected anthrax was sent to the DVO.

Following deaths an animal reported near Mburameizi Health center on September 14 laboratory samples were collected and sent to NADDEC. A public awareness drive was initiated on September 17, 2024. A provisional quarantine was instituted on September 18, 2024, prohibiting the movement, sale, or consumption of livestock and related products within, into, or out of Kanungu District. The MoH deployed the NRRT on September 22, 2024, and the district launched a mass livestock vaccination of susceptible livestock against anthrax on September 23, 2024. *Bacillus anthracis* was later confirmed on October 7, 2024.

#### Human health

From interaction with previously affected community members, the earliest human symptom onset was on June 27, 2024, in an adult male who had a history of participation in the slaughter and distribution of meat from cattle that had died suddenly on farm X. He later sought treatment at a local drug shop on July 11, 2024.

On September 9, 2024, the first human case was reported at Mburameizi Barracks HC III, and a notification was promptly sent to the DHO. The case-patient presented with characteristic anthrax skin lesions following exposure to meat from animals that died suddenly, and this triggered a notification to the DHO Laboratory. Samples were collected on September 14 and sent to the Uganda Virus Research Institute (UVRI) for testing. On September 17, 2024, UVRI confirmed two samples positive for *Bacillus anthracis* (Fig. 2).

**Figure 2.**
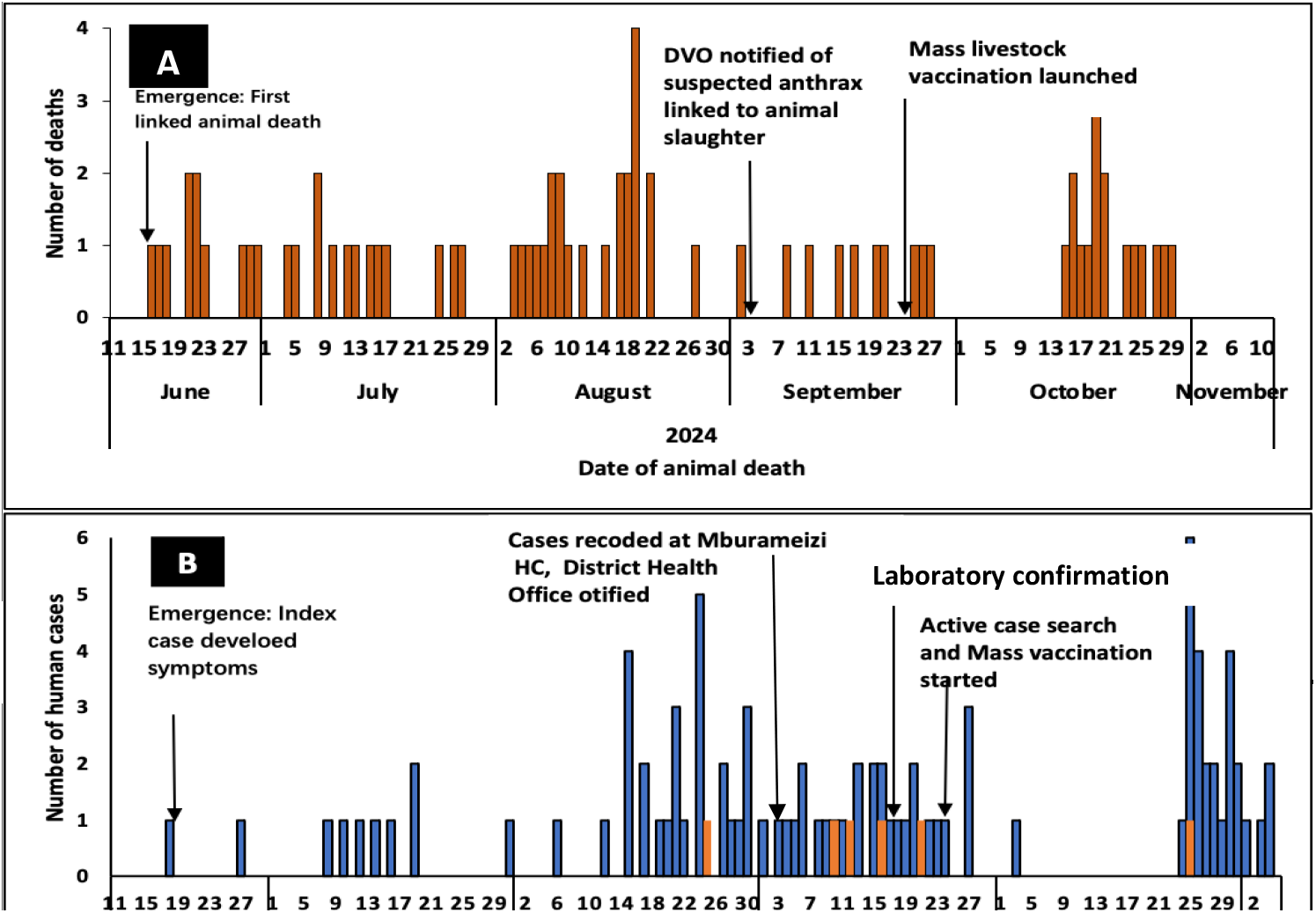
Epicurve of animal deaths (Panel A), human cases and human deaths (Panel B) during an anthrax outbreak in Kanungu District, June – November 2024.

On the same day, September 17, 2024, district authorities launched an awareness campaign by radio. The Ministry of Health deployed the National Rapid Response Team (NRRT) on September 22, 2024, to support outbreak investigation and response efforts. The investigation started with a records review and an active case search on September 23, 2024. (Fig. 2).

One Health response activities continued, with active case search and mass livestock vaccination against anthrax starting on September 24, 2024 (Table 2).

**Table 2:**
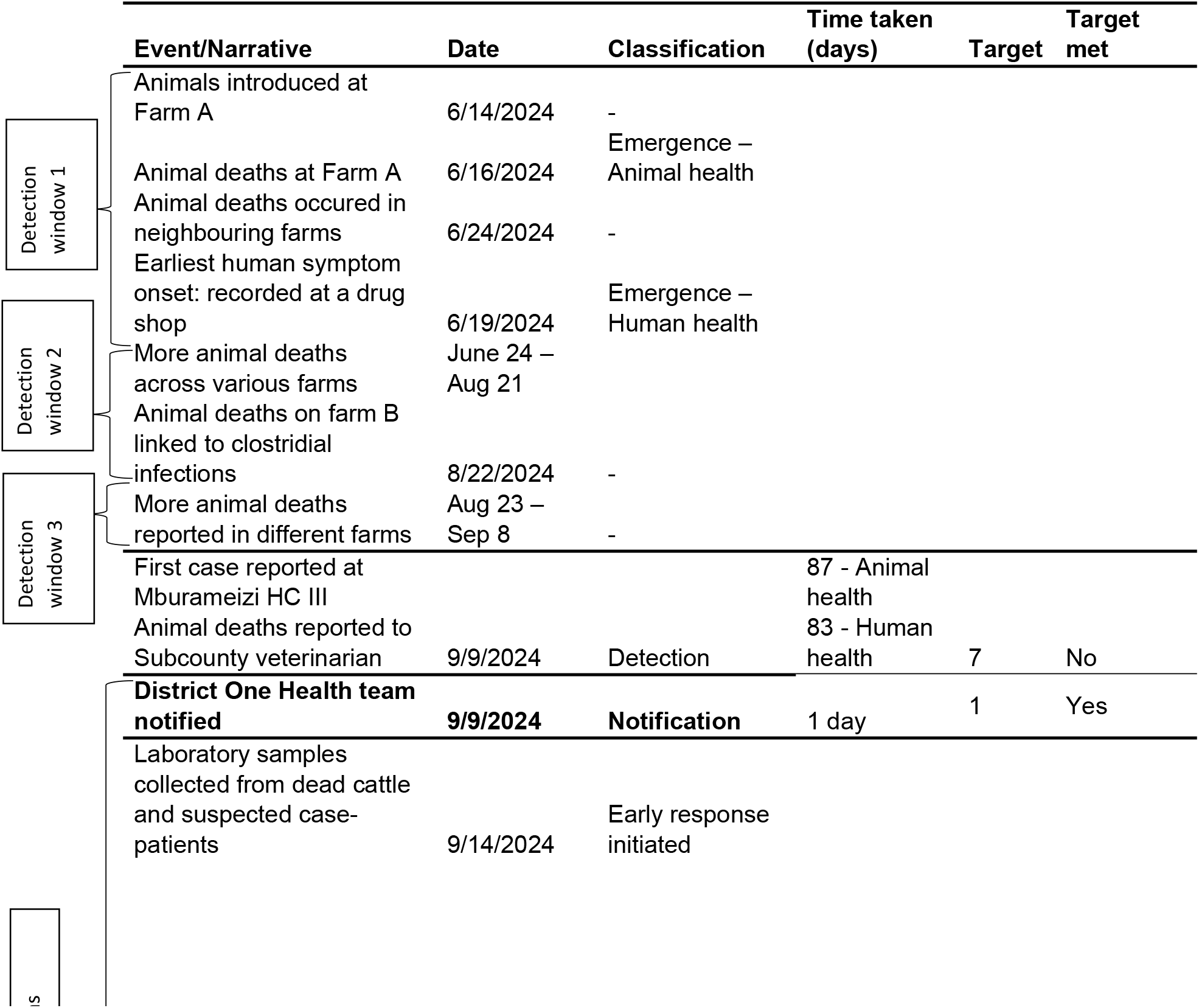
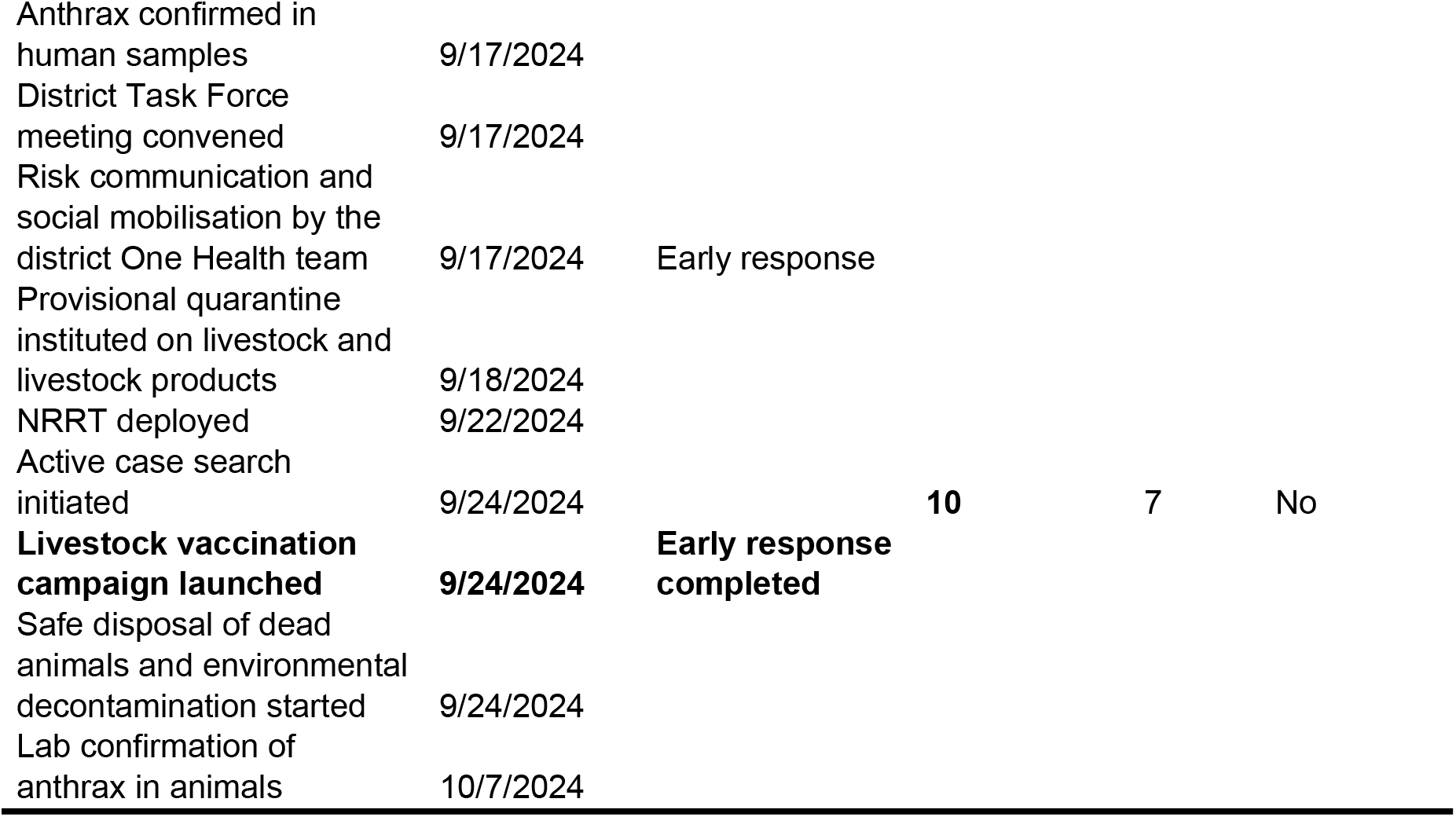
Key events and dates during an anthrax outbreak in Kanungu district, Southwestern Uganda, September-October 2024.

### Qualitative assessment

#### Animal Health

From engagements with different livestock owners, case-persons, health workers, and district political, technical, and administrative leaders, we documented bottlenecks and enablers for detection, notification, and response (Table 3).

**Table 3:** Bottlenecks and enablers for timely detection and response for the anthrax outbreak in Kanungu District, October 2024.

| Interval | Bottlenecks<br>Factors that prevented timely action. | Enablers<br>Factors that enabled timely action. |
| --- | --- | --- |
| Detection | 1. Community unawareness of anthrax, being the first known outbreak in decades. Most community members had never seen anthrax in their lifetime<br>2. Ready market for meat from dead animals within the community and in the DRC<br>3. Low suspicion index for anthrax among health workers<br>4. Service interruptions in the 6767 electronic 6767 platform | - |
|  | 5. Weak supervision of private health facilities. Many private health facilities were managing case-patients without referring them to government facilities, notifying or reporting to district authorities<br>6. |  |
| Notification | 1. Community practices: consumption of meat from dead animals was a well-known and widespread behaviour in the district | A real-time communication platform (WhatsApp) was available for health workers. This enabled rapid information sharing following detection and onward transmission to district authorities. |
| Response | 1. Human resource shortages, particularly in animal health. At the time of the outbreak, the most affected subcounty lacked a stationed animal health worker, hindering response detection and response efforts at the community level.<br>2. The District One Health Team had minimal prior exposure in coordinating response to zoonotic disease outbreaks, this being the first documented zoonotic incident in the district.<br>3. Logistical challenges in the NADDEC. At the time, the NADDEC did not have adequate laboratory supplies to analyse animal samples and confirm anthrax quickly. | 1. The district mobilised support from various partners, including international organisations as well as local private not-for-profit health organisations, as part of local coordination and resource mobilisation<br>2. Capacity building at the community level: Village Health Teams (VHTs) were trained to enhance their ability to detect and report suspected cases within their community.<br>3. Training animal health personnel: Animal health workers received targeted training on clinical recognition and reporting of anthrax and other animal diseases<br>4. Availability of anthrax vaccines from both MAAIF and on the open market |

To better understand how these enablers and bottlenecks influenced outbreak detection, notification and response, we collected some verbatim quotes aligned to detection, notification and response in animal and human health.

#### Detection

Interviews with stakeholders revealed that many respondents had encountered anthrax for the first time, which contributed to its delayed identification within the surveillance system. Several animal health workers reported a low suspicion index for anthrax, mainly due to the absence of prior field experience with similar cases. This was echoed in the reflections shared by some frontline animal health workers: “*I studied anthrax in school, but I have never seen any animal that has died of anthrax”. (Farm manager-)*.

*Animals have been dying for a long time, and they are usually healthy-looking animals. In such cases, we typically suspect Black Quarter. In fact, around August, some farmers vaccinated their herds against Black Quarter”. (Animal Health worker)*.

#### Notification

Notification was sent to the DVO by the district surveillance Focal Person (DSFP) for human health, after obtaining information from a WhatsApp platform for health workers. A notification in human health prompted a similar notification in animal health.

#### Response

The response was initiated along with the district health team. This started with public awareness on anthrax through radio talk shows, and announcement of a provisional quarantine on movement, sale and consumption of livestock and livestock products. The animal health team was unable to independently initiate and sustain response activities, owing to logistical and staffing challenges in the district animal health sector. One anima health worker had this to say:

*“We are not many animal health workers, not enough to cover all the sub-counties. It is difficult to follow up on animal health issues in such a wide area”. – Animal Health Worker*

Additionally, investigations into animal deaths were initiated, and mass livestock vaccination targeting the most affected subcounties was launched.

### Assessment in Human Health

#### Detection

One of the patients had a history of livestock rearing and involvement in slaughter and meat trade from animals that had died suddenly, raising suspicion of a possible anthrax outbreak. This delayed recognition in humans was ultimately what triggered suspicion of anthrax in livestock as well. Reflecting limited local awareness, one case person remarked: *“I used to hear about anthrax from old people, but at my age, I had never seen anyone with the disease*.*”* (*Case-patient)*

### Notification

The district maintains a dedicated WhatsApp group for disease surveillance, where health workers from various facilities share real-time information on emerging health issues. This system enabled prompt communication and action once the suspected case was identified. One health worker emphasized the utility of this platform: *“We have a WhatsApp group where different health workers all over the district share information. This helps us to know what is happening in different facilities in real time*.*” (Health worker)*.

### Response

Once the district team received laboratory results confirming *Bacillus anthracis*, an anthrax outbreak was announced. A joint One Health risk communication campaign was launched immediately, including radio talk shows to raise public awareness. At the same time, working with the animal health team, the district announced a provisional quarantine, prohibiting movement, sale, and consumption of all livestock and related products. The local response activities were beefed up by the deployment of the NRRT to support the district team with an epidemiological investigation and response coordination (Table 1). One health worker remarked on the awareness of the severity of anthrax that prompted immediate joint response: *“Anthrax is a serious disease that can cause major disruption in community livelihoods. Once it was confirmed, we had to act fast*.*” (Health worker)*.

## DISCUSSION

In this evaluation of anthrax response in a rural border district using the 7-1-7 framework, we report critical delays in detection and early response action in both human and animal outbreaks. Detection occurred 83 days after symptom onset in humans and 87 days after the first suspected animal deaths, both far exceeding the seven-day benchmark. In contrast, notification was achieved within one day, enabled by an established One Health communication platform. Early response actions were jointly initiated within 5 days of notification, and completed within 10 days, slightly exceeding the target. The outbreak, which resulted in 111 animal deaths and 90 human cases, including six fatalities, highlighted both systemic weaknesses, including limited frontline capacity for zoonotic surveillance and poor engagement of private facilities, and strengths, such as real-time communication and rapid intersectoral coordination once the outbreak was identified. Overall, these findings underscore the importance of improving early detection capacity and sustaining cross-sector collaboration to enhance national readiness for zoonotic threats.

Application of 7-1-7 to this outbreak revealed three critical windows for early detection missed by the surveillance system. First, an illegally imported animal into a private farm died exhibiting classic signs suggestive of anthrax, including the absence of rigor mortis and the presence of non-clotting blood oozing from natural orifices, features well-documented in anthrax-infected carcasses [1]. Despite these clear indicators, the carcasses were slaughtered, and the meat was distributed within the local community and to commercial meat dealers, with no notification to animal health workers. In this community, the consumption of meat from animals that had died suddenly was a normalized practice, as previous experiences had not resulted in any widely recognized health crises. Consequently, this behaviour persisted, with communities unaware of the broader implications. The continued occurrence of unexplained animal deaths in this area throughout August and September 2024 represents another critical phase for possible early detection. During this period, the animal deaths were reported to animal health workers, but were clinically misdiagnosed as Black Quarter. Animal health workers in service at the time had previously not seen a typical case of anthrax, as no such outbreaks had been previously reported in the district. Third, our evaluation found that livestock grazing and movement in the district were largely unregulated, which likely facilitated the undetected spread of anthrax to multiple subcounties, which likely enabled movement of exposed animals to different areas in the district, with subsequent animal deaths, slaughter and meat distribution facilitating spread of anthrax, leading to detection of the first human case. The farms in the affected areas were typically unfenced, allowing cattle to move freely across farms. This aligns with previous studies highlighting the role of uncontrolled animal movement in spreading zoonotic diseases during an active outbreak [23]. Combined, these gaps show how weak surveillance systems in animal health can negatively impact the timely detection of zoonotic threats in human health. Governments should invest in frontline animal health surveillance capacity and continued risk communication and engagement to promote awareness and timely reporting.

In addition to the missed detection opportunities, several human cases presented to community-level health facilities with hallmark signs of cutaneous anthrax—swelling, itching, and the formation of eschars. However, these symptoms did not raise any suspicion among healthcare providers or community members. This reflects limited clinical exposure and inadequate training among health workers to easily recognize anthrax, an infectious and often fatal zoonotic disease. Its endemicity in such areas with sporadic outbreaks demands sustained attention by governments. Similar findings on weaknesses in clinical recognition have been reported in rural settings across multiple countries in SSA, where endemicity and absence of prior outbreaks led to low suspicion for anthrax, even when symptoms were consistent with clinical definitions [14, 24-26]. This underscores the urgent need for ongoing health worker refresher training to maintain a high suspicion index, as well as continuous investment and improvement in diagnostic capacity to ensure the timely identification and reporting of zoonotic threats worldwide.

A significant contributing factor to this delayed response was the shortage of veterinary personnel in the district. Notably, the initially affected subcounty lacked any stationed veterinary staff at the time of the outbreak. It was only after cases began emerging that an assistant veterinary officer from a neighboring subcounty was temporarily assigned to support the affected communities. Prior studies have emphasized the pivotal role of veterinary professionals in public health and zoonotic disease detection and response, especially in rural low-resource settings [27-31]. Still, this critical workforce gap likely contributed to delays in both detection and formal reporting in this outbreak. Such shortages of trained animal health workers and uneven deployment to communities have been documented in LMICs, especially in rural areas where human-animal interaction is more frequent [32, 33]. In contrast, settings with stronger veterinary infrastructure, such as in Europe, Australia, and the USA, tend to detect zoonotic threats more quickly owing to comparatively fair distribution of trained animal health workers and integrated reporting strategies. These differences highlight the need for a dual approach. First, governments should prioritize training, recruitment, and rational deployment, especially in high-risk and underserved areas. Second, there should be efforts to strengthen community-based surveillance with emphasis on One Health. This double-pronged approach leverages both technical capacity and community engagement to enhance One Health preparedness and response systems.

Moreover, the outbreak was compounded by the existence of a ready market for meat from animals that had died suddenly. Following a surge in cattle deaths, the local prices of meat drastically reduced, and this economic incentive likely fueled continued slaughter and distribution of potentially infected carcasses, contributing to further silent spread of the disease. Meat was sold not only within local communities and townships but also across the border in the Democratic Republic of Congo (DRC). Such practices are not uncommon in sub-Saharan Africa, where communities frequently consume meat from animals that die suddenly, even during ongoing anthrax outbreaks [34]. Notably, the informal cross-border trade poses significant regional and transboundary threats to health and trade, and undermines ongoing public health measures. Findings from this evaluation highlight the need for governments to enforce stringent regulation of the movement of livestock and livestock products, and further highlight the need for community education on safe meat practices to prevent similar outbreaks in the future.

The absence of a functional, cost-free reporting system at the community level limited chances of early detection through the community-based Village Health Teams (VHTs). The guidelines for integrated disease surveillance and reporting (IDSR) provide for strengthening surveillance at the community level [35]. In Uganda, this was operationalized through the establishment of electronic event-based surveillance hotlines and reporting platforms, such as sending short messaging services (SMS) to 6767 [36]. Despite its introduction in the district and training, the local digital reporting platform had not worked in over two years. As shown in previous studies, VHTs can only be as effective as the surveillance and reporting tools available to them [37]. The community-level reporting gap in this outbreak highlights the need for regular system checks and dedicated focal persons to supervise and monitor community-level health reporting, ensuring access to and proper functionality of available surveillance and reporting tools. Conversely, a district-wide WhatsApp group enabled timely communication among health workers, facilitating rapid notification once the outbreak was detected. While informal, such platforms have proven helpful in public health surveillance and could be leveraged alongside formal systems, as documented elsewhere in different settings [38, 39].

Overall, our assessment revealed a delayed response timeline of 66-1-10 days in the human health sector and 85-1-14 days in the animal health sector, falling short of the recommended 7-1-7 benchmark. Despite having trained Village Health Teams (VHTs), weak coordination with the limited veterinary workforce hindered early detection. The District One Health Team and leadership lacked prior experience managing large-scale zoonotic outbreaks and were unprepared to implement a coordinated One Health response. As a result, three critical early detection windows were missed, highlighting the need for well-trained, adequately staffed, and coordinated One Health teams at the district level to enhance early detection and response to zoonotic threats [40].

Our findings underscore several public health and policy implications geared explicitly toward improving detection and response to zoonotic outbreaks. First, the delays in identifying both animal and human cases highlight the need to strengthen frontline surveillance systems. This can be achieved by addressing staffing gaps, particularly among veterinary personnel, and ensuring that community-based actors such as community health workers are empowered with functional reporting tools. Regular maintenance and monitoring of these systems should be institutionalized to ensure their reliability during outbreaks. Second, integrating the 7-1-7 framework into animal health surveillance, alongside human health, could promote early recognition and synchronised response to zoonotic events. Operationalizing this approach requires formal guidance, district-level orientation, and inter-sectoral simulation exercises to test readiness [19, 20]. Third, the demonstrated value of real-time digital communication among health workers calls for its formalisation and scale-up across districts. Establishing district-level One Health coordination mechanisms with routine information-sharing protocols could ensure swift activation and collaboration during health emergencies. Lastly, engaging private-sector actors, such as local drug sellers, meat vendors, and private veterinary practitioners in formal surveillance channels could bridge current detection gaps, especially in remote and underserved areas.

### Study limitations

Our study has limitations. First, our qualitative findings primarily relied on participants’ ability to recall events that had occurred three months prior. This may have introduced inaccuracies, particularly regarding dates, case counts, and the interpretation of specific events. To enhance reliability, multiple members of the investigation team independently reviewed and analysed the recorded interviews. Additionally, we validated the information through triangulation, utilizing insights from district leadership and comparing it with data collected from outbreak investigation and response activities. Second, the dynamics of transmission and response likely differ between districts and countries, and the results may not be generalizable. However, the results reflect a prototypical outbreak in a prototypical agrarian setting and provide valuable learnings that could be replicated in similar settings. Finally, the 7-1-7 framework has not yet been formally adopted within Uganda’s animal health surveillance and outbreak response system. We aimed to explore and demonstrate the practical application, relevance and utility of the 7-1-7 framework within a typical One-Health context, as a simple, actionable tool with potential to strengthen coordination among One Health sectors during routine disease surveillance and outbreak response.

### Conclusion

This anthrax outbreak response revealed critical delays in detection despite timely notification and relatively prompt early response once the outbreak was identified. The evaluation further revealed strengths in cross-sectoral coordination but highlighted weaknesses in zoonotic disease surveillance and frontline detection capacity. Strengthening animal health surveillance, building private sector engagement, and training frontline workers to recognize zoonoses early could improve future outbreak responsiveness.

## Data Availability

The data upon which our findings are based belong to the Uganda Public Health Fellowship Program, and can be made available upon reasonable request from the corresponding author, subject to permission from the Uganda Public Health Fellowship Program.

https://717alliance.org/

## List of abbreviations

BINP: Bwindi Impenetrable National Park
DHO: District Health Officer
DHT: District Health Team
DLG: District Local Government
DOHT: District One Health Team
DRC: Democratic Republic of Congo
DTF: District Task Force
DVO: District One Health Officer
HC: Health Centre
MoH: Ministry of Health
NADDEC: National Animal Diseases Diagnostics and Epidemiology Centre
NRRT: National Rapid Response Team
PEPFAR: Presidential Emergency Plan for
AIDS: Relief
QENP: Queen Elizabeth National Park
RDC: Resident District Commissioner
VHT: Village Health Teams

## Funding and disclaimer

This study was supported by the President’s Emergency Plan for AIDS Relief (PEPFAR) through the United States Centers for Disease Control and Prevention Cooperative Agreement number GH001353-01 through Makerere University School of Public Health to the Uganda Public Health Fellowship Program, Ministry of Health. The contents of this manuscript are solely the responsibility of the authors and do not necessarily represent the official views of the US Centers for Disease Control and Prevention and the Department of Health and Human Services, Makerere University School of Public Health, or the Uganda Ministry of Health.

## Acknowledgements

We acknowledge the generous support and tireless dedication of the Kanungu District Local Government technical team, led by the District Health Team, who provided continuous technical support to the 7-1-7 assessment and the general outbreak response. We also acknowledge the district political leadership led by the Resident District Commissioner (RDC), who was also the head of the District Task Force (DTF). The RDC continuously supported risk communication and implementation of control measures that eventually led to the end of the outbreak. Special appreciation goes to Dr Scott Kellerman and the entire team at Bwindi Community Hospital, Buhoma, for the time you dedicated to participating in the outbreak investigation and this assessment, including availing your vehicle and fuel. Finally, we acknowledge the team at the Global Health Security Department of the Infectious Diseases Institute that provided continuous mentorship and technical guidance during the period of this study.

## Author contribution

HK led the conception, design, analysis, and interpretation of the study results and drafted the manuscript. HK, CM, BA, AT, and BMR participated in the outbreak investigation, conception, design, analysis, and interpretation of results. MB, ME, GKN, PJL, SAA, SA, LA, analysed the laboratory samples and participated in the analysis and interpretation of the results, and reviewed the manuscript for intellectual content. RM and HNN supervised the outbreak investigation and reviewed the manuscript for intellectual content. HTN, BK, LN, JJL, PK, DMB, FK, LB, and ARA reviewed the manuscript draft for intellectual content and scientific integrity. All authors reviewed the final manuscript draft.

## Conflict of Interest

The authors declared no conflict of Interest.

## Notes

### Competing Interest Statement

The authors have declared no competing interest.

### Author Declarations

This was an early action review conducted during a public health emergence. As such, the Uganda Ministry of Health waives requirements for IRB approvals. The MoH authorised this study, and the office of the Center for Global Health, US Center for Disease Control and Prevention determined that this activity was not human subject research, with its primary intent being for public health practice or disease control. This activity was reviewed by CDC and was conducted consistent with applicable federal law and CDC policy.

## References

1. Galante, D. and A. Fasanella. Anthrax in Animals. 2022 2022/07 [cited 2022; Available from: https://www.msdvetmanual.com/infectious-diseases/anthrax/anthrax-in-animals.

2. Organization, W.H., International health regulations (2005). 2008: World Health Organization.

3. Index, G.H.S. What is the GHS Index? 2021 [cited 2025 July 10, 2025]; Available from: https://ghsindex.org/about/#What-is-the-GHS-Index.

4. Organization, W.H., International Health Regulations (2005). Third edition. 2016, Geneva: World Health Organization.

5. Ario, A.R., et al., Uganda’s experience in establishing an electronic compendium for public health emergencies. PLOS Global Public Health, 2023. 3(2): p. e0001402.

6. Kayiwa, J., et al., Establishing a public health emergency operations center in an outbreak-prone country: lessons learned in Uganda, January 2014 to December 2021. Health security, 2022. 20(5): p. 394–407.

7. Kabasa, J.D., J. Kirsten, and I. Minde, Implications of changing agri-food system structure for agricultural education and training in Sub-Saharan Africa. Journal of Agribusiness in Developing and Emerging Economies, 2015. 5(2): p. 190–199.

8. Faostat, F., New food balances. FAOSTAT. Available via FAO. Accessed, 2021. 25.

9. Statistics, U.B.o., NATIONAL LIVESTOCK CENSUS 2021: MAIN REPORT. 2024, Government of Uganda: Kampala, Uganda.

10. Mbonye, A.K., et al., Ebola viral hemorrhagic disease outbreak in West Africa-lessons from Uganda. African health sciences, 2014. 14(3): p. 495–501.

11. Buregyeya, E., et al., Operationalizing the one health approach in Uganda: challenges and opportunities. Journal of epidemiology and global health, 2020. 10(4): p. 250–257.

12. Frieden, T.R., et al., 7-1-7: an organising principle, target, and accountability metric to make the world safer from pandemics. The Lancet, 2021. 398(10300): p. 638–640.

13. Bochner, A.F., et al., Implementation of the 7-1-7 target for detection, notification, and response to public health threats in five countries: a retrospective, observational study. The Lancet Global Health, 2023. 11(6): p. e871–e879.

14. Bush, L.M. and M.T. Vazquez-Pertejo. Anthrax. 2023 2023/03 [cited 2023; Available from: https://www.msdmanuals.com/professional/infectious-diseases/gram-positive-bacilli/anthrax.

15. Nuwamanya, Y., et al., Anthrax outbreaks in western Uganda: The role of illegal meat dealers in spreading the infection, August 2022–April.

16. Monje, F., et al., Animal Anthrax outbreak triggered by butchering infected carcasses on and or near the pastureland, Kween District, Uganda: January–December 2018.

17. Monje, F., et al., Animal Anthrax outbreak caused by slaughtering infected carcasses on and or near the pastureland, Kiruhura District, Uganda, May-October 2018.

18. Wafula, M.M., A. Patrick, and T. Charles, Managing the 2004/05 anthrax outbreak in queen elizabeth and lake mburo national parks, uganda. African Journal of Ecology, 2008. 46(1): p. 24–31.

19. Nantima, N., et al., The importance of a One Health approach for prioritising zoonotic diseases to focus on capacity-building efforts in Uganda. Rev Sci Tech, 2019. 38(1): p. 315–25.

20. Sekamatte, M., et al., Multisectoral prioritization of zoonotic diseases in Uganda, 2017: A One Health perspective. PloS one, 2018. 13(5): p. e0196799.

21. Medley, A.M., et al., Preventing the cross-border spread of zoonotic diseases: Multisectoral community engagement to characterize animal mobility—Uganda, 2020. Zoonoses and Public Health, 2021. 68(7): p. 747–759.

22. Lydia, N., et al., Implementing the 7-1-7 target to improve epidemic preparedness and response in Uganda. BMJ Global Health, 2025. 10(7): p. e018207.

23. Hasahya, E., et al., Analysis of patterns of livestock movements in the Cattle Corridor of Uganda for risk-based surveillance of infectious diseases. Frontiers in Veterinary Science, 2023. 10: p. 1095293.

24. Kungu, J.M., et al., Perceptions and practices towards anthrax in selected agricultural communities in Arua District, Uganda. Journal of Tropical Medicine, 2020. 2020(1): p. 9083615.

25. Hamutyinei Dhliwayo, T., et al., Anthrax outbreak investigation in Tengwe, Mashonaland West Province, Zimbabwe, 2022. PLoS One, 2022. 17(12): p. e0278537.

26. Tumusiime, L., et al., Anthrax outbreak linked to consumption and handling of meat from unexpectedly deceased cattle, Kyotera district, Uganda, June–December 2023. One Health Outlook, 2025. 7(1): p. 1–9.

27. Ilukor, J., et al. Analysis of veterinary service delivery in Uganda: an application of the process net-map tool. in Proceedings of the Tropentag 2012 conference: Resilience of agricultural systems against crises. 2012.

28. Ilukor, J., et al., THE PROVISION OF VETERINARY SERVICES: WHO ARE THE INFLUENTIAL ACTORS AND WHAT ARE THE GOVERNANCE CHALLENGES? A CASE STUDY OF UGANDA. Experimental Agriculture, 2015. 51(3): p. 408–434.

29. Shrivastava, P., S.K. Sahoo, and K. Venkatesh, Role of Veterinarians in Public Health. Epidemiology and Environmental Hygiene in Veterinary Public Health, 2025: p. 287–298.

30. Braam, D.H., et al., A fair share? Animal health actors and resources in One Health initiatives: A multisite case study in Ethiopia and Pakistan. CABI One Health, 2023(2023): p. ohcs202300015.

31. Worsley-Tonks, K.E.L., et al., Strengthening global health security by improving disease surveillance in remote rural areas of low-income and middle-income countries. Lancet Glob Health, 2022. 10(4): p. e579–e584.

32. Getachew, A., et al., Association between domestic animal exposure and diarrhea prevalence in under-five children in low- and middle-income countries: a systematic review and meta-analysis. BMC Pediatrics, 2024. 24(1): p. 601.

33. Rubenstein, B.L., et al., Community-based Guinea worm surveillance in Chad: Evaluating a system at the intersection of human and animal disease. PLoS neglected tropical diseases, 2021. 15(3): p. e0009285.

34. Kock, R., et al., A One-Health lens for anthrax. The Lancet Planetary Health, 2019. 3(7): p. e285–e286.

35. Organization, W.H., Technical Guidelines for Integrated Disease Surveillance and Response in the African Region October 2010, in Technical Guidelines for Integrated Disease Surveillance and Response in the African Region October 2010. 2010.

36. Organization, W.H., Integrated disease surveillance in the African region: a regional strategy for communicable diseases 1999-2003, in Integrated disease surveillance in the African Region: a regional strategy for communicable diseases 1999-2003. 1999. p. 24–24.

37. Sanou, H., et al., How community-based health workers fulfil their roles in epidemic disease surveillance: a case study from Burkina Faso. BMC Health Services Research, 2024. 24(1): p. 1372.

38. Adesina, T., et al., Social Media and disease surveillance in Nigeria–the Role of WhatsApp. Annals of Global Health, 2017. 83(1): p. 19–20.

39. Carrillo, M.A., et al., WhatsApp-based intervention in urban Colombia to support the prevention of arboviral diseases: a feasibility study. Pathogens and Global Health, 2024. 118(4): p. 334–347.

40. Walekhwa, A.W., et al., Strengthening anthrax outbreak response and preparedness: simulation and stakeholder education in Namisindwa district, Uganda. BMC Veterinary Research, 2024. 20(1): p. 484.

